# Effect of co-infection with parasites on severity of COVID-19

**DOI:** 10.1101/2021.02.02.21250995

**Authors:** Teklay Gebrecherkos, Zekarias Gessesse, Yazezew Kebede, Atsbeha Gebreegzabher, Geremew Tasew, Mahmud Abdulkader, Hiluf Ebuy, Abraham Desta, Atakilti Hailu, Vanessa Harris, Tobias Rinke de Wit, Dawit Wolday

## Abstract

**Background:** Severe acute respiratory syndrome coronavirus 2 (SARS-CoV-2) infection results in a spectrum of clinical presentations. The effect of co-infection with parasites on the clinical features of COVID-19 is unknown.

**Methods:** We prospectively enrolled consecutive COVID-19 patients and screened them for intestinal parasitic infections. Patients were followed during hospitalization for clinical outcomes. Patients with parasitic co-infection were compared to those without parasitic co-infection. The primary outcome was the proportion of COVID-19 patients who developed severe disease. Factors associated with the development of severe disease were determined by logistic regression.

**Results:** A total of 515 patients with PCR-confirmed SARS-CoV-2 infection were screened for intestinal parasites, of whom 267 (51.8%) were co-infected with one or more parasites. Parasitic co-infection correlated inversely with COVID-19 severity. Severe COVID-19 was significantly higher in patients without parasites [47/248 (19.0%, CI: 14.52-24.35)] than in those with parasites [21/267 (7.9%, CI: 5.17-11.79)]; p<0.0001. There was a significantly higher proportion of patients who developed severe COVID-19 in the non-protozoa group [56/369 (15.2%, CI: 11.85-19.23)] as compared to the protozoa group [12/146 (8.2%, CI: 4.70-14.00)]; p=0.036. Significant higher proportion of the patients presented at baseline with severe COVID-19 in the helminth negative group [57/341 (16.7%, CI: 13.10 – 21.08)] than in the group with pre-existing helminth infection [11/174 (6.3%, CI: 3.51 – 11.11)]; p=0.001. In addition, after adjustment for age and presence of comorbidities, COVID-19 patients with any parasite co-infection [aOR 0.41 (95% CI: 0.22–0.77); p=0.006], or with protozoa co-infection [aOR 0.45 (95% CI: 0.21–0.98); p=0.044] as well as those with helminth co-infection [aOR 0.37 (95% CI: 0.17–0.80); p=0.011] had lower probability of developing severe COVID-19 compared with those without parasite, protozoa or helminth co-infection.

**Conclusion:** Our results suggest that co-infection with parasitic co-infection appears to be associated with reduced COVID-19 severity. The results suggest that parasite-driven immunomodulatory responses may mute hyperinflammation associated with severe COVID-19.

## Introduction

Infection with severe acute respiratory syndrome coronavirus 2 (SARS-CoV-2) results in a spectrum of clinical presentations. Whereas most people with COVID-19 develop only asymptomatic or mild illness, at-risk patients can develop severe disease requiring hospitalization and respiratory [1-3]. In severe cases, COVID-19 can be complicated by the acute respiratory distress syndrome (ARDS), sepsis and septic shock, multi-organ failure, including acute kidney injury and cardiac injury [2-4]. Older age and underlying comorbidities due to non-communicable diseases (NCDs) such as hypertension, cardiovascular diseases and diabetes, have been reported as risk factors for disease severity and death [4-8].

Underlying conditions in the setting of low and medium-income countries (LMICs) are different from those in high-income countries (HICs). In LMICs, infectious diseases are highly prevalent than NCDs. One such infectious diseases are neglected infectious disease (NIDs). Most notably, parasitic infections affect more than 2 billion people throughout the world, with disproportionately high prevalence rates in resource-poor settings [9, 10]. Multicellular and highly complex parasites such as *Ascaris*, hook worm, *Trichuris, Enterobius* and *Schistosoma*, as well as unicellular organisms including *Entamoeba, Giardia, Toxoplasma, Cyclospora* and *Cryptosporidia* are among the major organisms that contribute to the global intestinal parasitic disease burden [9, 10]. The distribution of parasitic infections varies widely in the different parts of the world.

Chronic and persistent parasitic infection is common in LIMCs, and chronic parasitic infections, possibly in part through direct modulation of the host’s immune responses, have been shown to alter clinical outcomes to other infections [11, 12]. Pre-existing parasitic infections may also modify the host’s immune response to infection with SARS-CoV-2 with possible beneficial or detrimental effects [13-19]. Recent reports demonstrated an inverse correlation between the incidence of COVID-19 and with soil-transmitted helminths, schistosomiasis or malaria [20]. To the best of our knowledge, however, there have been no studies to date that have assessed the effect of co-infection with parasites on the severity of COVID-19. In this study, therefore, we compared the clinical outcomes of COVID-19 patients with or without parasitic co-infection.

## Methods

### Study design and participants

We identified individuals recruited to the Profile-CoV study who had been screened for parasitic infections. Profile-CoV project (Clinicaltrials.gov: NCT04473365) is a prospective observational cohort study being undertaken in two sites in Ethiopia, with the aim of profiling of immunological response to SARS-CoV-2 in the context of persistent immune activation in Sub-Saharan Africa.

All patients suspected of having SARS-CoV-2 infection were screened for SARS-CoV-2 infection with a nasopharyngeal real-time polymerase chain reaction (RT-PCR). All patients with confirmed SARS-CoV-2 infection were admitted into Kuyha Isolation Hospital run by Mekelle University College of Health Sciences, Mekelle, Northern part of Ethiopia for isolation and treatment. Patients were admitted irrespective of the clinical severity status. Admitted patients receive supportive therapy according to clinical need. Patients with severe disease receive high-flow oxygen via nasal cannula or intubation as well as dexamethasone. All COVID-19 cases confirmed by PCR test result between July and October 2020 were included in this study.

Sociodemographic, clinical and laboratory data were collected using standardized Case Record Forms (CRFs) adapted from the International Severe Acute Respiratory and Emerging Infection Consortium’s (ISARIC) CRFs for emerging severe acute respiratory infections [21]. Patient’s clinical status was stratified following WHO criteria as asymptomatic, mild/moderate, severe (with dyspnea, respiratory rate ≥ 30 breaths per minute, O2 saturation ≤ 93%, lung infiltrates ≥ 50% of the lung fields within 24-48 hours), and critical (with respiratory failure, septic shock, and/or multiple organ failure) [22]. All data were then entered onto electronic medical records.

### Laboratory analysis

SARS-CoV-2 infection was confirmed by RT-PCR on samples obtained from nasopharyngeal swabs, according to WHO guidelines. Fresh stool sample specimens were obtained for examination for ova and parasites. Analysis included direct microscopic examination and modified Ritchie concentration method [23]. In addition, the intensity of infection was determined using Kato-Katz method and was calculated and reported as individuals’ eggs per gram of feces (EPG), as described previously [23, 24] and recommended by the WHO [25]. Helminth-positive individuals were stratified into three infection intensity categories: (i) light (1– 99 EPG), (ii) moderate (100–399 EPG), and (iii) heavy (≥400 EPG).

### Ethical considerations

Patients enrolled provided written consent to participate in to the Profile-CoV study. The study protocol was reviewed and approved by the Health Research Ethics Review Committee (HRE RC) of Mekelle University College of Health Sciences (#ERC 1769/2020). All individual identifiers were de-linked from the original sources.

### Statistical analysis

The primary outcome for this study was the proportion of COVID-19 patients who developed severe disease among patients with and without a parasitic co-infection. Asymptomatic and mild/moderate cases were classified as non-severe cases and both severe and critical were classified as severe cases. Baseline characteristics for continuous variables were expressed as the median with interquartile range (IQR), and for categorical variables as proportions. Whereas categorical variables were compared using χ^2^ test or Fisher’s exact test, categorical variables were compared by Mann-Whitney U or Kruskal-Wallis tests, as appropriate. The association between parasitic co-infection and COVID-19 severity was determined by logistic regression analysis. Independent variables, including age, sex, comorbidity, parasite infection (overall and disaggregated by parasite type into protozoa and helminths), were included in the initial univariate analysis. Then a multivariate regression analyses [adjusted odds ratio (aOR)] were calculated (with backward stepwise elimination) by including all variables that were p<0.20 by univariate analysis. P values <0.05 were considered statistically significant. Data was analyzed using STATA (Statistical package v. 14.0, StataCorp, Texas, USA).

## Results

Baseline socio-demographic data of the study participants is summarized in Table 1. The majority of our study population were male (62.5%). The median age of the cohort was 32 (IQR 26–43) years, the majority (60.7%) being in the age range 24 to 44 years. Most notably, 86.8% the cohort population were either asymptomatic or had mild/moderate symptoms at the time of diagnosis. The remaining (13.2%) had either severe disease or required admission to intensive care unit (ICU). COVID-19 patients with severe disease tended to be older and had had significantly higher symptoms, including cough (82.4% vs. 21.9%), dyspnea (70.6% vs. 2.9%), fever (61.8% vs. 13.9%), head ache (41.2% vs. 15.4%), chest pain (25.0% vs. 1.1%), myalgia (25.0% vs. 3.1%), sore throat (20.6% vs. 4.3%), hemoptysis (13.2% vs. 1.6%) and diarrhea (5.9% vs. 1.3%), when compared to those presenting with non-severe form of COVID-19.

**Table 1.** Clinical features among COVID-19 patients without or with parasitic co-infection

| Characteristic | All patients<br>N=515 | Without parasite<br>N= 248 | With parasite<br>N= 267 | P value |
| --- | --- | --- | --- | --- |
| **Socio-demographic features:** |  |  |  |  |
| Gender, N (%) |  |  |  |  |
| Female | 193 (37.5) | 90 (36.3) | 103 (38.6) | 0.592 |
| Male | 322 (62.5) | 158 (63.7) | 164 (61.4) |  |
| Age in years [median (IQR)] | 32 (26-43) | 33 (26-47) | 32 (25-41) | 0.064 |
| Age group [years, N (%)] |  |  |  |  |
| < 24 | 79 (15.4) | 34 (13.8) | 45 (16.9) | 0.203 |
| 24 – 44 | 312 (60.7) | 145 (58.7) | 167 (62.6) |  |
| 45 – 59 | 77 (15.0) | 40 (16.2) | 37 (13.9) |  |
| ≥ 60 | 46 (9.0) | 28 (11.3) | 18 (6.7) |  |
| **Clinical symptoms and signs:** |  |  |  |  |
| Fever, N (%) | 104 (20.2) | 62 (25.0) | 42 (15.7) | 0.009 |
| Dyspnea, N (%) | 61 (11.9) | 45 (18.2) | 16 (6.0) | <0.0001 |
| Cough (any type, N (%)) | 154 (29.9) | 84 (33.9) | 70 (26.2) | 0.058 |
| Non-productive cough | 109 (21.2) | 57 (23.0) | 52 (19.5) |  |
| Productive cough | 45 (8.7) | 27 (10.9) | 18 (6.7) |  |
| Hemoptysis, N (%) | 16 (3.1) | 14 (5.7) | 2 (0.8) | 0.001 |
| Chest pain, N (%) | 22 (3.7) | 15 (6.1) | 7 (2.6) | 0.055 |
| Sore throat, N (%) | 33 (6.4) | 20 (8.1) | 13 (4.9) | 0.139 |
| Head ache, N (%) | 97 (18.8) | 60 (24.2) | 37 (13.9) | 0.003 |
| Nasal congestion, N (%) | 28 (5.4) | 12 (4.8) | 16 (6.0) | 0.564 |
| Loss of smell and/or taste, N (%) | 49 (9.5) | 26 (10.5) | 23 (8.6) | 0.470 |
| Diarrhea, N (%) | 10 (1.9) | 5 (2.0) | 5 (1.9) | 0.906 |
| Myalgia, N (%) | 31 (6.0) | 20 (8.1) | 11 (4.1) | 0.060 |
| Fever > 37.3 °C, N (%) | 18 (3.5) | 10 (4.0) | 8 (3.0) | 0.522 |
| Temperature (median °C, IQR) | 36.0 (36.0-36.7) | 36.0 (36.0-36.8) | 36.0 (36-36.7) | 0.114 |
| Systolic blood pressure (median mmHg, IQR) | 115 (108-125) | 115 (105-125) | 115 (110-124) | 0.747 |
| Diastolic blood pressure (median mmHg, IQR) | 75 (70-80) | 75 (70-80) | 75 (68-80) | 0.920 |
| Respiratory rate (median breaths/minute, IQR) | 22 (20-23) | 22 (20-24) | 22 (19-23) | 0.084 |
| Heart rate (median beats/minute, IQR) | 83 (75-90) | 85 (75-90) | 82 (75-90) | 0.3060 |
| **Comorbidities** |  |  |  |  |
| Comorbidity (at least 1) | 78 (15.2) | 49 (19.8) | 29 (10.9) | 0.005 |
| Non-communicable disease (NCDs) comorbidities | 61 (11.8) | 38 (15.3) | 23 (8.6) | 0.019 |
| Diabetes | 31 (6.0) | 22 (8.9) | 9 (3.4) | 0.009 |
| Hypertension | 28 (5.4) | 16 (6.5) | 12 (4.5) | 0.328 |
| Cardio-vascular diseases | 5 (1.0) | 4 (1.6) | 1 (0.4) | 0.152 |
| Chronic obstructive lung diseases and asthma | 14 (2.7) | 10 (4.0) | 4 (1.5) | 0.077 |
| Chronic liver disease | 3 (0.6) | 3 (1.2) | 0 (0.0) | 0.071 |
| Chronic kidney disease | 7 (1.4) | 5 (2.0) | 2 (0.8) | 0.215 |
| Communicable disease comorbidities |  |  |  |  |
| HIV | 8 (1.6) | 5 (2.0) | 3 (1.1) | 0.410 |
| Tuberculosis | 1 (0.2) | 1 (0.4) | 0 (0.0) | 0.298 |
| **Outcomes** |  |  |  |  |
| Admission to ICU | 29 (5.6) | 20 (8.1) | 9 (3.4) | 0.021 |
| Supplemental oxygen | 68 (13.2) | 47 (19.0) | 21 (7.9) | <0.0001 |
| Invasive mechanical ventilation | 13 (2.5) | 9 (3.6) | 4 (1.5) | 0.123 |
| Death | 3 (0.6) | 3 (1.2) | 0 (0.0) | 0.071 |
| Severe COVID-19 | 68 (13.2) | 47 (19.0) | 21 (7.9) | <0.0001 |

Of the total 515 individuals enrolled in the study, 267 (51.8%) harbored one or more intestinal parasites (Table 2). Protozoa and helminth infections comprised 28.4% and 33.8%, respectively. The most common protozoa infections were *Entamoeba spp*. (23.5%) and *Giardia* (4.6%). Among helminths, the most common infections were *H. nana* (21.2%), *S. mansoni* (6.2%), and *A. lumbricoides* (4.9%). There was no difference in gender and age distribution when those with parasites were compared with those without parasites (Table 2). However, those without parasite co-infection presented with significant higher proportion of fever (25.0% vs. 15.7%), head ache (24.2% vs. 13.9%), dyspnea (18.2% vs 6.0%) and hemoptysis (5.7% vs. 0.8%) than those with parasite co-infection. In addition, the proportion of comorbid conditions, in particular NCDs, were significantly higher in COVID-19 patients without parasitic co-infection when compared to those with parasitic co-infection (Table 1).

**Table 2.** Clinical features and prevalence of parasitic infections among non-severe and severe COVID-19 patients

| Parasite co-infection | All patients<br>N=515 | Non-severe patients<br>(asymptomatic and<br>mild/moderate)<br>N= 447 | Severe patients<br>(severe and critical)<br>N= 68 | P value |
| --- | --- | --- | --- | --- |
| Any parasite | 267 (51.8) | 246 (55.0) | 21 (30.9) | <0.0001 |
| Protozoa – all | 146 (28.4) | 134 (30.0) | 12 (17.7) | 0.036 |
| - *Entamoeba histolytica/dispar* cyst | 104 (20.2) | 98 (21.9) | 6 (8.8) | 0.012 |
| - *Entamoeba histolytica* trophozoite | 17 (3.3) | 15 (3.4) | 2 (2.9) | 0.859 |
| - *Giardia lamblia* cyst | 8 (1.7) | 8 (1.8) | 2 (2.9) | 0.521 |
| - *Giardia lamblia* trophozoite | 15 (2.9) | 13 (2.9) | 2 (2.9) | 0.988 |
| Helminth – all | 174 (33.8) | 163 (36.5) | 11 (16.2) | 0.001 |
| - *Hymenolopis nana* | 109 (21.2) | 103 (23.0) | 6 (8.8) | 0.007 |
| - *Schistosoma mansoni* | 32 (6.2) | 30 (6.7) | 2 (2.9) | 0.230 |
| - *Ascaris lumbricoides* | 25 (4.9) | 22 (4.9) | 3 (4.4) | 0.855 |
| - *Trichuris trichiura* | 11 (2.1) | 11 (2.5) | 0 (0.0) | 0.191 |
| - Hook worm | 9 (1.8) | 9 (2.0) | 0 (0.0) | 0.238 |
| - *Taenia* spp. | 2 (0.4) | 2 (0.5) | 0 (0.0) | 0.580 |
| - Soil-transmitted helminths only | 44 (8.5) | 41 (9.2) | 3 (4.4) | 0.191 |
| Poly-parasitism – any | 65 (12.7) | 62 (13.9) | 3 (4.4) | 0.028 |
| - Protozoa plus helminth | 50 (9.7) | 48 (10.8) | 2 (2.9) | 0.043 |
| - Helminth plus helminth | 17 (3.3) | 17 (3.8) | 0 (0.0) | 0.102 |
| **Outcomes** |  |  |  |  |
| Admission to ICU | 29 (5.6) | 1 (0.2) | 28 (41.2) | <0.0001 |
| Supplemental oxygen | 68 (13.2) | 1 (0.2) | 67 (98.5) | <0.0001 |
| Invasive mechanical ventilation | 13 (2.5) | 0 (0.0) | 13 (19.1) | <0.0001 |
| Death | 3 (0.6) | 0 (0.0) | 3 (4.4) | <0.0001 |

The proportion of COVID-19 patients with parasitic infection decreased with increasing categories of disease severity, in particular for any parasitic and helminthic infections (Figure 1). In addition, clinical outcomes were considerably worse in COVID-19 patients without parasites (Table 1). Of all COVID-19 patients, a higher proportion of patients without parasitic co-infection were admitted to ICU and required supplemental oxygen compared to those with parasitic co-infection (Table 1). Severe COVID-19 was significantly higher in patients without parasites [47/248 (19.0%, CI: 14.52-24.35)] than in those with parasites [21/267 (7.9%, CI: 5.17-11.79)]; p<0.0001. There was a significantly higher proportion of patients who developed severe COVID-19 in the non-protozoa group [56/369 (15.2%, CI: 11.85-19.23)] as compared to the protozoa group [12/146 (8.2%, CI: 4.70-14.00)]; p=0.036. Significant higher proportion of the patients presented at baseline with severe COVID-19 in the helminth negative group [57/341 (16.7%, CI: 13.10 – 21.08)] than in the group with pre-existing helminth infection [11/174 (6.3%, CI: 3.51 – 11.11)]; p=0.001. However, we did not observe any correlation between helminth egg-load with COVID-19 severity.

**Figure 1.**
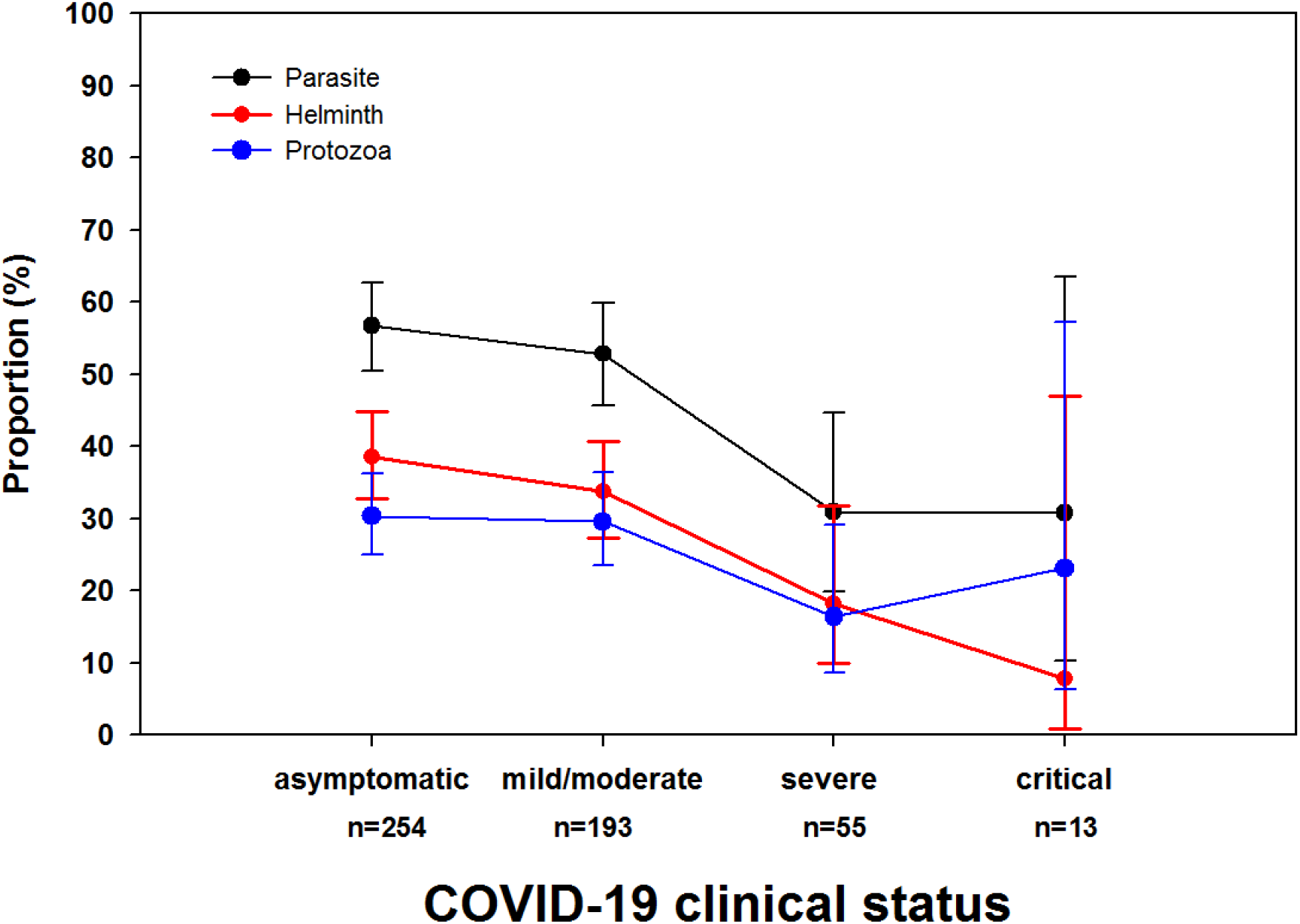
Proportion of parasites, protozoa and helminths among COVID-19 patients with asymptomatic, mild/moderate, severe and critical clinical presentation. P-values for trend (p=0.002, p=0.006 and p=0.196 for any parasite, helminth and protozoa, respectively).

In univariate analysis, older age, diabetes, hypertension, cardio-vascular diseases, chronic obstructive lung diseases, chronic liver disease, and chronic kidney disease were all associated with increased likelihood of COVID-19 severity (Table 3). In the contrary, having any parasite, protozoal infection and helminth infections were all associated with reduced probability of severe COVID-19. In multivariate analysis, older age, hypertension and chronic kidney diseases were associated with the odds of COVID-19 severity. In addition, after adjustment for age and presence of comorbidities, COVID-19 patients with any parasite co-infection [aOR 0.41 (95% CI: 0.22–0.77); p=0.006; Table 3], or with protozoa co-infection [aOR 0.45 (95% CI: 0.21–0.98); p=0.044] as well as those with helminth co-infection [aOR 0.37 (95% CI: 0.17–0.80); p=0.011] had lower probability of developing severe COVID-19 compared with those without parasite, protozoa or helminth co-infection. Interestingly, we noted that the odds of having a NCD was significantly lower in COVID-19 patients having co-infection with parasites (OR: 0.52 95% CI: 0.30-90.2, p=0.020), or helminths (OR 0.26 95% CI: 0.12-0.56; p=0.001).

**Table 3.**
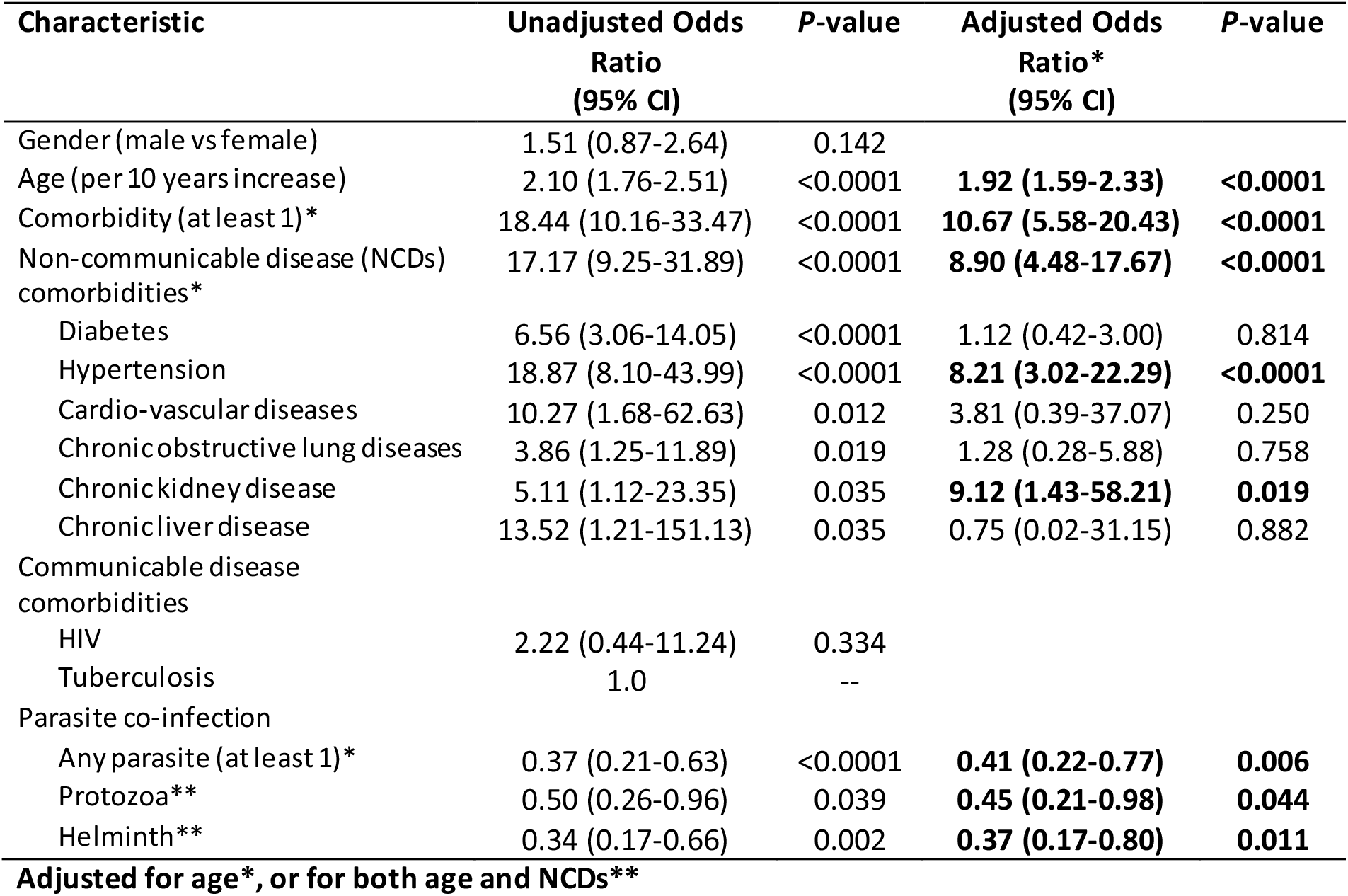
Factors associated with severity among COVID-19 patients in Northern Ethiopia

## Discussion

This study demonstrated that co-infection with enteric parasites, both protozoa and helminths, was associated with lower probability of developing severe COVID-19. To the best of our knowledge, the findings represent the first report regarding an association between parasites and COVID-19 severity. Notably, the association was maintained even after adjusting for age and presence of comorbid conditions, factors commonly associated with COVID-19 severity.

In addition, previous reports have demonstrated that helminthic infection reduces the risk of development of diabetes and metabolic syndrome in humans [26-34]. This is in line with our finding that pre-existing chronic co-infection with parasites/helminths in our cohort was associated with a significant lower proportion and reduced risk of NCDs. This may suggest that the observed decreased COVID-19 severity in our cohort might be attributed through decreased NCD risk.

The pathogenesis of severe disease in COVID-19 has been linked to the phenomenon of immune hyperactivation [35, 36], that resembles that of chronic inflammatory condition, such as hypertension, obesity, diabetes and inflammatory bowel diseases [37-41]. It is possible, therefore, that parasites mute COVID-19 severity through their effects in modulating systemic immune response. Parasitic infections modulate human’s immune systems, in such a way that the T-helper (TH) responses are augmented along with predominant regulatory (Treg) responses [11]. This parasite-driven TH2 response, in turn, may counterbalances TH1 responses, responsible for the hyperinflammation associated with severe COVID-19 [11]. In addition, parasite-driven gut microbiome changes may modulate the host’s immune response [12]. Thus, it is possible that parasitic infections may affect pathogenesis both through direct modulation of the immune system as well as through parasite-driven microbiome balance. Indeed, it has been demonstrated in animal models that enteric helminth protect against pulmonary viral infections through interaction with microbiota [42].

The strengths of the current study include the prospective nature of the study design. However, our study has some limitations. First, stool samples were not able to be collected for every consecutive patient which may have resulted in a potential selection bias. Second, stool examination was determined by microscopy only. The presence of very low intensity parasite infection determined by PCR, though shown to be superior to microscopy with increased sensitivity and specificity [43], might indeed preclude the effects on immune modulation to have any significant effect. Third, the effect of individual parasite species on COVID-19 severity could not be ascertained in the current study because of the small sample size in groups with different parasite species. Finally, the inclusion of smaller proportion of severe cases as compared to non-severe cases in our cohort may potentially bias the results.

In conclusion, our study demonstrates for the first time that pre-existing parasitic infection, both with protozoa and helminths, may provide some sort of protection from the pathology linked with severe COVID-19. This is corroborated by the observed low fatality rate of COVID-19 in LMICs settings where parasitic infections are endemic [15, 18, 20, 44]. We also recommend to expand the study to other LMICs and also include the effect of the interplay between parasite-microbiome on COVID-19 severity. Unraveling the mechanisms underlying severe COVID-19 offers avenues for novel preventive and therapeutic interventions.

## Data Availability

Data are available upon reasonable request. All requests to access the data must be processed through the project Research Steering Committee. All data accessibility requests should be directed to the corresponding authors.

## Acknowledgement

We thank all the staff for their contribution and the study participants for their cooperation. This research was supported by grants from the European and Developing Countries Clinical Trials Partnership (EDCTP), supported by the European Union (RIA-2020EF-2095) and Joep Lange Institute for Global Health and Development, The Netherlands.

## Notes

### Competing Interest Statement

The authors have declared no competing interest.

### Clinical Trial

NCT04473365

### Author Declarations

The study protocol was reviewed and approved by the Health Research Ethics Review Committee (HRERC) of Mekelle University College of Health Sciences (#ERC 1769/2020).

